# A Pooled Electronic Consultation Program to Improve Access to Genetics Specialists

**DOI:** 10.1101/2023.02.08.23284667

**Authors:** Emma K. Folkerts, Renée C. Pelletier, Daniel C. Chung, Susan A. Goldstein, Douglas S. Micalizzi, Kristen M. Shannon, David A. Sweetser, Eugene K. Wong, Heidi L. Rehm, Leland E. Hull

## Abstract

Innovative service delivery models are needed to increase access to genetics specialists. Electronic consultation (e-Consult) programs can connect clinicians with specialists. At Massachusetts General Hospital, an e-Consult service was created to address genomics-related questions. In its first year, the e-Consult service triaged 153 requests and completed 122 in an average of 3.2 days. Of the 95 e-Consults with actionable recommendations, there was documentation that most ordering clinicians followed through (82%). A variety of providers used the service, although the majority (77%) were generalists. E-Consult models should be considered as one way to increase access to genetics care.

## Introduction

Access to genetic subspecialists for care-related questions is often limited.^1–3^ Electronic consultations (**e-Consults**) have been trialed across diverse health systems to provide expedited access to specialists.^4–6^ e-Consult programs within genetics and genomics programs care have emerged as models to address access issues by providing first-pass triage of genetics questions, answering simple questions that do not require a visit (thus freeing access for additional patient care), as well as educating generalists on genetics care topics that could be directly applied to future patients.

Massachusetts General Hospital (MGH) is an academic medical center in Boston, MA, where several specialties offer clinical genetics care.^7^ In April 2021, we launched a pooled e-Consult program to provide timely access to subspecialists from three different clinics, The MGH Center for Cancer Risk Assessment, MGH Medical Genetics and Metabolism, and the MGH Preventive Genomics Clinic. Clinicians order an e-Consult through the electronic health record (**EHR)**, select a reason for the e-Consult from a prespecified list, and enter a patient-specific clinical question with the option to add additional data or relevant reports. A genetic counseling assistant performs initial triage of submitted e-Consults by directing inquiries to a specific specialty in the “pool” of genetics specialists based on the clinical content. The answering specialist reviews the patients’ EHR data and sends an electronic response to the submitting clinician, which is then documented in the EMR. Sometimes, a genetic counselor or another staff member may investigate the submitted topic and draft a response. E-Consults can be declined at any point in the process.

We characterized the patients and clinicians utilizing the e-Consult program and the e-Consult processes during the program’s first year.

## Methods

We performed a retrospective observational analysis of e-Consult data that the Mass General Brigham Institutional Review Board approved.

We characterized the providers ordering, the providers responding to, and the patients for whom e-Consults were ordered between April 2021 and March 2022, as well as the content of and outcomes of e-Consults that were collected over this period. We obtained our data from 1) the Mass General Physicians Organization (MGPO) e-Consult database, which pulls relevant data from the EHR, 2) publicly available MGH websites characterizing providers and their clinics, and 3) chart review. We characterized patients (age at order, legal sex, and self-reported race), providers submitting e-Consults (degree and specialty), and providers responding to e-Consults (specialty). We characterized e-Consults by several factors: turnaround time, the prespecified reason for the e-Consult, e-Consult recommendation type (actionable v. no action recommended), and recommendation follow-through by the submitting provider (Yes vs. No/Not Documented).

Patient and provider demographics were summarized descriptively. Next, we outlined the reasons for e-Consult submission and the outcomes of e-Consults.

## Results

Of 153 e-Consults submitted in the program’s first year, 122 were completed (80%), and 31 were declined (Figure 1). The reasons for declined e-Consults are summarized in Figure 1. A minority of e-Consults related to patients over age 65 (8%); patients were primarily female and White (**Table 1**). Both physicians and nurse practitioners submitted e-Consults; a greater proportion of nurse practitioner e-Consults were declined (39%) compared to physician-submitted e-Consults (18%). Clinicians requesting e-Consults represented a large range of specialties, although the majority (80/104, 77%) were generalists such as internists, general pediatricians, and family medicine physicians. Most e-Consults were triaged to Medical Genetics and Metabolism (90/122, 73%), followed by Cancer Genetics (n=31, 25%). All clinicians answering e-consults were physicians.

**Table 1.**
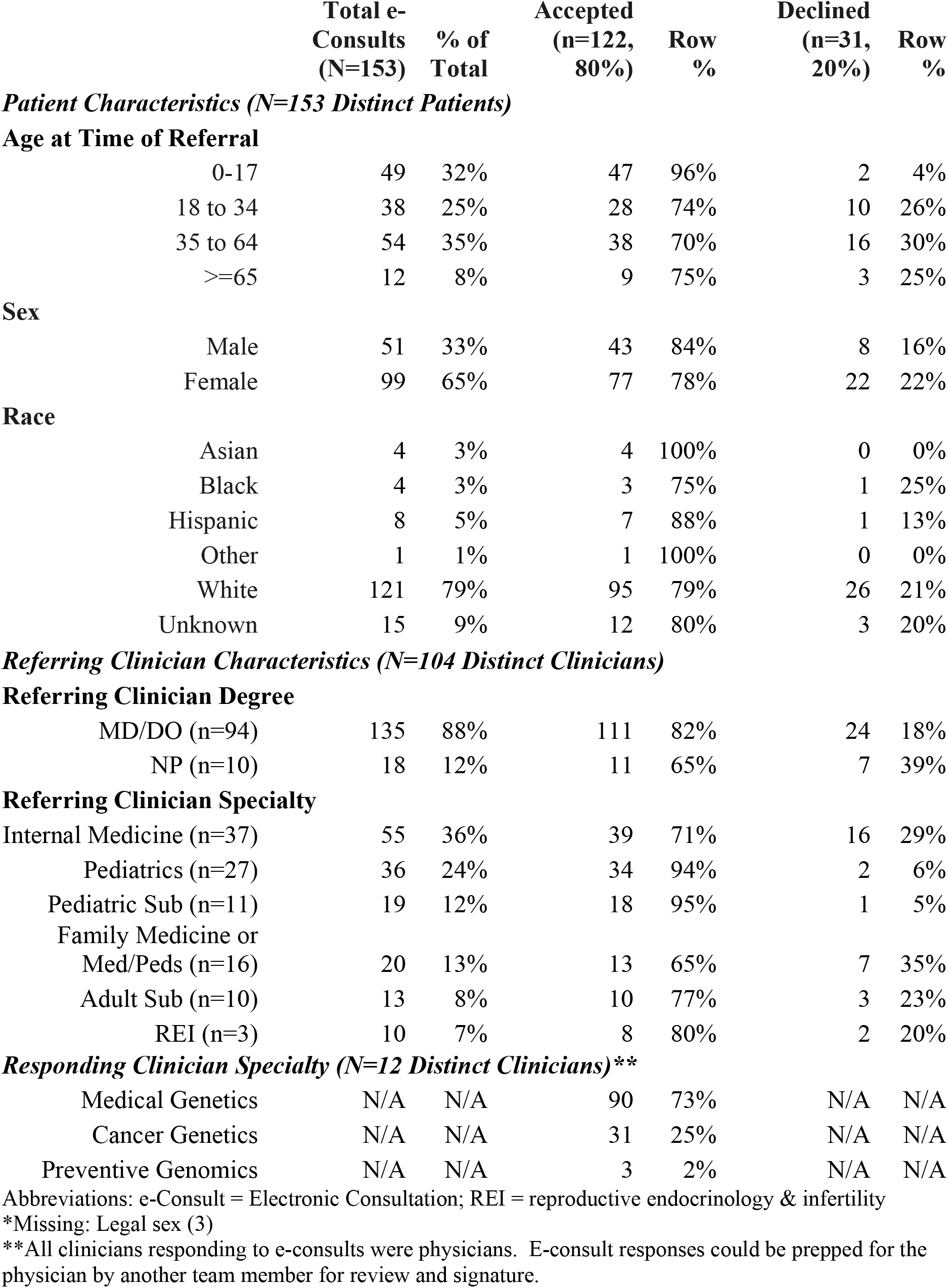
Characteristics of e-Consults Ordered (N=153 distinct e-Consults)

|  | Total e-Consults<br>(N=153) | % of<br>Total | Accepted<br>(n=122,<br>80%) | Row<br>% | Declined<br>(n=31,<br>20%) | Row<br>% |
| --- | --- | --- | --- | --- | --- | --- |
| ***Patient Characteristics (N=153 Distinct Patients)*** |  |  |  |  |  |  |
| **Age at Time of Referral** |  |  |  |  |  |  |
| 0-17 | 49 | 32% | 47 | 96% | 2 | 4% |
| 18 to 34 | 38 | 25% | 28 | 74% | 10 | 26% |
| 35 to 64 | 54 | 35% | 38 | 70% | 16 | 30% |
| >=65 | 12 | 8% | 9 | 75% | 3 | 25% |
| **Sex** |  |  |  |  |  |  |
| Male | 51 | 33% | 43 | 84% | 8 | 16% |
| Female | 99 | 65% | 77 | 78% | 22 | 22% |
| **Race** |  |  |  |  |  |  |
| Asian | 4 | 3% | 4 | 100% | 0 | 0% |
| Black | 4 | 3% | 3 | 75% | 1 | 25% |
| Hispanic | 8 | 5% | 7 | 88% | 1 | 13% |
| Other | 1 | 1% | 1 | 100% | 0 | 0% |
| White | 121 | 79% | 95 | 79% | 26 | 21% |
| Unknown | 15 | 9% | 12 | 80% | 3 | 20% |
| ***Referring Clinician Characteristics (N=104 Distinct Clinicians)*** |  |  |  |  |  |  |
| **Referring Clinician Degree** |  |  |  |  |  |  |
| MD/DO (n=94) | 135 | 88% | 111 | 82% | 24 | 18% |
| NP (n=10) | 18 | 12% | 11 | 65% | 7 | 39% |
| **Referring Clinician Specialty** |  |  |  |  |  |  |
| Internal Medicine (n=37) | 55 | 36% | 39 | 71% | 16 | 29% |
| Pediatrics (n=27) | 36 | 24% | 34 | 94% | 2 | 6% |
| Pediatric Sub (n=11) | 19 | 12% | 18 | 95% | 1 | 5% |
| Family Medicine or<br>Med/Peds (n=16) | 20 | 13% | 13 | 65% | 7 | 35% |
| Adult Sub (n=10) | 13 | 8% | 10 | 77% | 3 | 23% |
| REI (n=3) | 10 | 7% | 8 | 80% | 2 | 20% |
| ***Responding Clinician Specialty (N=12 Distinct Clinicians)\*\**** |  |  |  |  |  |  |
| Medical Genetics | N/A | N/A | 90 | 73% | N/A | N/A |
| Cancer Genetics | N/A | N/A | 31 | 25% | N/A | N/A |
| Preventive Genomics | N/A | N/A | 3 | 2% | N/A | N/A |

Abbreviations: e-Consult = Electronic Consultation; REI = reproductive endocrinology & infertility
\*Missing: Legal sex (3)
\*\*All clinicians responding to e-consults were physicians. E-consult responses could be prepped for the physician by another team member for review and signature.

**Figure 1.**
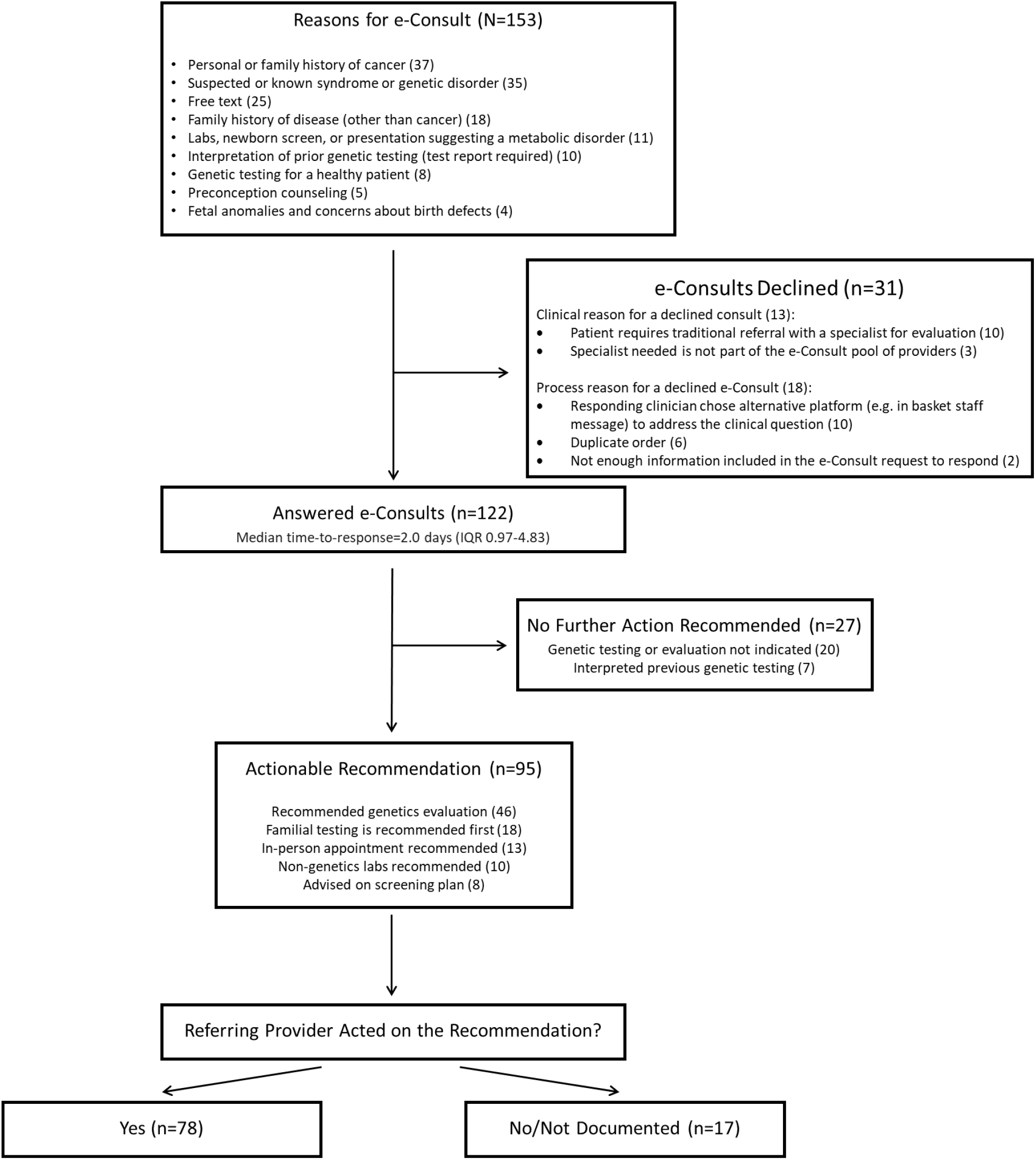
Outcomes of Submitted e-Consults.

The top pre-specified reasons ordering providers selected for e-Consult submission were a personal or family history of cancer (n=37), suspicion or knowledge of a known syndrome or genetic disorder (n=35), and reasons not captured by the other prespecified fields (free text, n=25) (**Figure 1**). E-Consults were answered with a median of 2.0 days. Most completed e-Consults resulted in at least one actionable recommendation for the submitting clinician (95/122); 82% (78/95) of those with an actionable recommendation had documented evidence of follow-through.

## Discussion

With increasing indications for incorporating genetics into patient care, building systems to improve access to genetics expertise is paramount. In the first year of our e-Consult program, diverse questions were submitted by clinicians from various specialties, especially generalist providers. As primary care clinicians have endorsed poor knowledge and comfort providing genetics care to their patients,^8,9^ the successful use of the e-Consult program by generalists is encouraging because such programs can provide a system for interacting and learning from genetics experts.

Our data suggest that an e-Consult program can help to reduce unnecessary visits. There were several potential sources of saved visits that we observed. For example, of the 122 e-Consults answered, 27 recommended no further action, 10 recommended non-genetics lab testing, and 8 advised screening plans. In addition, e-Consult recommendations can also improve the efficiency of subsequent genetics evaluations, such as when familial genetic testing was recommended first (18/122 answered e-Consults), to enhance the utility of subsequent genetics evaluations. Further studies should assess the impact of e-consults on access to genetics services across the hospital system.^10^

Finally, a significant number of e-Consults (20%) were declined. The primary reasons for declining were either clinical (e.g., appropriate specialist not in the e-Consult “pool” of providers) or process errors (e.g., duplicate order entries). To scale the program, we aim to resolve clinical barriers to using the program, such as recruiting additional specialists from fields of high demand to join the consult “pool,” refining workflows, and further educating referring providers on the appropriate use of the E-Consult program.

## Data Availability

The participants of this study did not give written consent for their data to be shared publicly, so due to the sensitive nature of the research supporting data is not readily available. Specific requests can be addressed to the corresponding author.

